# Course and severity of post-H1N1 narcolepsy type 1: a long-term prospective cohort study

**DOI:** 10.64898/2026.03.25.26349255

**Authors:** Kristin Langdalen, Louise Frøstrup Follin, Rannveig Viste, Janita Vevelstad, Ragnhild Berling Grande, Hilde T. Juvodden, Per M Thorsby, Jonas Gjesvik, Marte K. Viken, Ketil Størdal, Berit Hjelde Hansen, Stine Knudsen-Heier

## Abstract

**Objective:** The narcolepsy type 1 (NT1) phenotype severity is heterogeneous, and the disease course is largely unknown. The 2009–10 H1N1-(Pandemrix®)-vaccinations were followed by increased numbers of possibly more severe post-H1N1 NT1 cases but long-term prospective data on large, vaccinated cohorts are missing.

**Methods:** 130 consecutive post-H1N1 NT1 cases (113/130 Pandemrix®-vaccinated) were prospectively followed up after approximately 5.5 years. Epworth Sleepiness Scale (ESS), cataplexy, hypnagogic hallucinations, sleep paralysis, PSG, MSLT, and BMI were evaluated. Phenotype severity predictors (hypocretin-1 deficiency severity <40 vs. 40-150 pg/ml; Pandemrix®-vaccination; disease duration) were tested in age and sex-adjusted multivariable regressions.

**Results:** From baseline to follow-up, phenotype severity overall improved (milder symptoms, higher mean MSLT sleep latency (SL) and fewer SOREMPs, all p<0.001). Follow-up phenotype severity was strongly predicted by the same baseline measures. Females had worse ESS and cataplexy, men had higher BMI, and young individuals had lower mean MSLT SL and more SOREMPs. Severe hypocretin deficiency (<40 pg/ml) predicted baseline PSG SOREMPs and lower MSLT SL. Vaccinated individuals had more severe baseline PSG/MSLT measures but greater long-term symptom improvement, and vaccination no longer predicted PSG/MSLT severity at follow-up.

**Conclusion:** The best prognostic factor for long-term NT1 phenotype severity is the earlier phenotype severity. Hypocretin deficiency severity also predicts parts of short-term but not long-term phenotype severity. Pandemrix-vaccination is associated with initially more severe phenotype but larger long-term improvement i.e. a different clinical course than in unvaccinated NT1, although medication effects cannot be excluded. Our findings underscore heterogeneity in NT1 phenotype and disease trajectories.

**Statement of significance:** This large prospective study demonstrates that long-term NT1 phenotype severity is strongly explained by the corresponding earlier phenotype severity. Hypocretin deficiency severity also predicts some phenotype severity measures, supporting that the degree of hypocretin system dysfunction influences the phenotype also in post-H1N1 NT1. Likewise, we confirm that age and sex affect NT1 phenotype severity. However, we additionally find that Pandemrix-vaccination is associated with initially more severe phenotype measures that improve more over time than in unvaccinated NT1. Our findings underscore the heterogeneity in NT1 phenotype and disease trajectories, which can direct more tailored prognosis, counselling, and long-term management. Identification of prognostic markers (baseline severity, vaccination, hypocretin deficiency severity, younger age, female sex) further supports clinical management/follow-up stratification in NT1.

## Introduction

Narcolepsy type 1 (NT1) is a chronic neurological sleep disorder characterized by excessive daytime sleepiness (EDS) and cataplexy, frequently accompanied by hypnagogic hallucinations, sleep paralysis, and fragmented nocturnal sleep [1]. These features are largely attributed to loss of hypocretin (also called orexin), a neuropeptide central for sleep-wake and muscle tonus regulation [2–5]. Weight gain and elevated body mass index (BMI) are also common in NT1, likewise attributed to the role of hypocretin in appetite, energy expenditure and lipid metabolism [2,6–11]. Although the NT1 core features are well described, the longitudinal clinical course and the determinants of phenotype severity are less characterized, leaving clinicians uncertain about prognosis and long-term management [12].

NT1 is considered a multifactorial disorder where genetic susceptibility (such as human leucocyte antigen (HLA)) combined with environmental triggers lead to autoimmune-mediated loss or dysfunction of hypocretin-producing neurons [4,5,13–15]. Environmental exposures include infections [16,17] and notably, the 2009–10 H1N1-vaccinations with Pandemrix® were followed by a marked NT1 incidence increase in several European countries including Norway [18–25].

Although hypocretin deficiency explains many NT1 manifestations, phenotypic expression is heterogeneous [6]. Some individuals quickly develop severe NT1 with all symptoms, whereas others present more insidiously with a partial symptom set or milder symptoms [12]. Across NT1 studies, several factors are associated with greater phenotype severity including HLA-DQB1\**06:02*-positivity, presence of low cerebrospinal fluid (CSF) hypocretin-1 level, younger age at disease onset, and female sex [11,26–34]. Higher BMI is associated with the presence of hypocretin deficiency (low hypocretin-1 values i.e. < 1/3 of normal mean) compared to those with normal CSF hypocretin-1 values in one sporadic human CNS hypersomnia cohort [35]. In animal NT1 models with gradual hypocretin neuron loss, cataplexy and weight gain slowly begin to appear after 75%-90% neuron loss and weight gain significantly continues to increase after almost complete (90%-99%) neuron loss [10,36]. However, whether the severity of hypocretin deficiency (undetectable versus low, but detectable hypocretin-1 levels) plays a role in the human NT1 phenotype severity/variation is sparsely studied. Undetectable CSF hypocretin-1 levels were not associated with BMI in a sporadic (unvaccinated) NT1 cohort [27] or BMI in our previously reported post-H1N1 (mainly Pandemrix®-vaccinated) NT1 cohort [9]. Undetectable hypocretin levels did though predict low MSLT SL and higher number of MSLT SOREMPs in a small sporadic NT1 study (n=18) [37], more severe sleep fragmentation [38,39] and more PSG muscle activations during sleep [40] in our post-H1N1 NT1 studies. Direct comparisons between studies are limited by different definitions of “undetectable” hypocretin levels and variable phenotype measures.

The long-term clinical course of NT1 remains less characterized. Although individuals with NT1 can develop additional symptoms and/or worsening of symptoms over time, on a group level an overall attenuation of core symptom severity after disease onset is reported in several studies on sporadic NT1 [32,41]. However, EDS severity has also been found mainly stable [42] or higher with advancing age [33]. BMI has been reported to increase over time in some sporadic NT1 cohorts [41,43–45] and in one of these also later stabilise with prolonged disease [45]. Disrupted nocturnal sleep has consistently been shown to worsen with age in sporadic NT1 [12,33,41,44]. Because large prospective NT1 studies are scarce, factors associated with phenotype improvement or deterioration remain poorly defined. In a small study of sporadic NT1 (n=26), milder hypocretin deficiency (low, but detectable CSF hypocretin-1 levels) and no complaints of disturbed nighttime sleep at baseline were associated with later EDS improvement [42]. The variable phenotype presentation and interindividual variability may suggest that NT1 comprises phenotype subvariants [46].

A central, still debated question is whether Pandemrix®-vaccinated NT1 represents a (more severe) phenotype subvariant [20,23,40,47–49]. Early reports described Pandemrix®- vaccinated NT1 individuals as surprisingly young, with more abrupt disease onset, rapid early weight gain, and more fulminant cataplexy [20,50]. Subsequent cross-sectional comparative studies have reported inconsistent and generally none/modest differences compared to sporadic NT1. Some of these did though report isolated findings of shorter mean multiple sleep latency test (MSLT) sleep latency (SL) [49,51], more SOREMPs [22,51], more sleep fragmentation [51], longer PSG latency [52] and more cataplexy and weight gain [53] in vaccine-associated groups. Interpretation has been limited by relatively small sample sizes (the largest study included 69 Pandemrix®-vaccinated NT1 individuals [51]), incomplete availability of CSF hypocretin and genetic data, heterogeneous treatment status, and variable definitions of vaccine-relation (7-58% Pandemrix®-vaccination rates in the “sporadic”/non-vaccine-related groups [51–53]).

Notably, only a single longitudinal study of Pandemrix®-vaccinated NT1 individuals has been published (26 vaccine-related versus 25 non-vaccine-related individuals), finding that individuals with vaccine-related NT1 initially had more frequent cataplexy and lower BMI, but no difference in PSG/MSLT measures between groups [54]. At follow-up, there was no marked symptom difference between groups, but BMI had increased in the vaccine-related group. PSG/MSLT measurements were not available/conducted at follow-up. Hypocretin deficiency severity in vaccine-related (but not in non-vaccine-related) NT1 individuals correlated with more severe symptoms (EDS and cataplexy) [54].

We therefore conducted a long-term prospective observational study of a large Norwegian national cohort of consecutively included well-characterized post-H1N1 NT1 individuals (i.e. with a disease onset after October 2009) +/- Pandemrix®-vaccination aiming to 1) describe longitudinal changes in subjective and objective phenotype measures of post-H1N1 NT1, and 2) identify clinical, demographic and biological (HLA-DQB1*06:02, CSF hypocretin-1) predictors of phenotype severity and clinical course, with an additional focus on Pandemrix®-vaccinated and unvaccinated NT1 differences.

## Methods

### Study design and participants

153 post-H1N1 NT1 individuals with symptom onset after October 2009 (i.e. after the onset of the 2009–10 H1N1 pandemic/Pandemrix®-vaccinations) were consecutively included at baseline between February 2015 and June 2024 at the Unit for Brain Disorders, Department of Rare Disorders, Oslo University Hospital (previously: Norwegian centre of expertise for neurodevelopmental disorders and hypersomnias, NevSom), Oslo University Hospital. Diagnostics were done by ESRS certified somnologists (SKH, BHH) according to ICSD-3/ICSD-3-TR criteria [55]. Individuals were included regardless of Pandemrix®-vaccination status. Exclusion criteria were secondary narcolepsy or major/serious comorbidity precluding participation. 130 individuals subsequently completed a follow-up re-assessment visit between February 2022 and June 2024. Individuals lost to follow-up were: 19 individuals who declined/could not participate due to acute illness, work commitments, or pregnancy/childcare, and 4 individuals included at baseline in 2023–2024 i.e. considered too late for the present follow-up inclusion. Subsets of the baseline cohort have previously been published [9,38,40,48]. The study was part of our unit‘s national post-H1N1 narcolepsy advisory/follow-up assignment prompted/funded by the Norwegian Ministry of Health and Care Services. Approval by the Regional Committees for Medical and Health Research Ethics Southeast Norway (REK approval 2014/450) was obtained, and all individuals gave their written informed consent (parents consented for children <16 years).

### Procedures

Prior to baseline, individuals were instructed to pause all medication known to affect sleep/wake or narcolepsy symptoms for at least two weeks [55]. At follow-up, individuals continued their usual treatment. The visits (baseline and follow-up) included neurological and psychiatric examination, semi-structured interviews, and validated questionnaires. Lifetime occurrence and frequency of core narcolepsy symptoms were assessed using the Norwegian translation of the Stanford Sleep Questionnaire [56], which includes Epworth Sleepiness Scale (ESS); ESS ≥ 11/24 indicated EDS [57]. Sex was recorded as sex assigned at birth. Individuals underwent overnight PSG followed by MSLT consisting of five 30-minute nap opportunities at two-hour intervals. Missing PSG/MSLT recordings were: 3 at baseline (2 technically invalid, 1 individual declined PSG/MSLT-investigation); at follow-up another individual declined PSG/MSLT-investigation. A wrist actigraph (Actiwatch, Philips) was worn for two weeks before PSG/MSLT (missing data at baseline: 4 individuals had no actigraphy; at follow-up: 2 had no actigraphy; 2 had 7 days actigraphy, 2 had 10 days actigraphy). All individuals with missing data had typical NT1 with cataplexy, hypocretin deficiency and HLA-DQB1\**06:02* positivity. PSG/MSLT recordings were acquired with SOMNOscreen Plus and scored in 30-second epochs by ESRS-certified somnologist-technician (JV) according to AASM criteria [58]. Nocturnal sleep fragmentation was assessed by sleep stage shift index ((SSSI) calculated as number of stage transitions per hour of total sleep time) and awakening index ((AI) calculated as number of awakenings per hour of total sleep time). Height, weight, and BMI (kg/m2) were measured/calculated. For children we computed age-and sex-adjusted BMI z-scores and classified weight status using the International Obesity Task Force thresholds corresponding to adult BMI cut-offs [59]. For adults, we applied standard adult BMI categories: normal weight 18.5–24.9, overweight 25.0–29.9, obese ≥30.0 kg/m2) [60]. Individuals <18 years at baseline were BMI analysed in the “children” group at both baseline and follow-up, and individuals ≥18 years at baseline were BMI analysed in the “adult” group. Lumbar puncture was performed prior to/after baseline inclusion at local hospitals and CSF hypocretin-1 was analysed at the Hormone Laboratory, Dept. of Medical Biochemistry, Oslo University Hospital. Hypocretin deficiency was defined by the international standard (< 1/3 of the normal laboratory mean; in Norway < 150 pg/ml) [61]. In a few cases, where two hypocretin measurements existed the one closest to baseline inclusion was used. Pandemrix®-vaccination status was obtained from the Norwegian Immunization Registry (SYSVAK). Pandemrix® was the only known H1N1-vaccine used in the present cohort. Thirteen individuals were not registered in SYSVAK but were included in the vaccinated group based on self-/parent-report on vaccination at school or workplace.

## Statistical analysis

Analyses were performed using Stata/SE 18. Normality of continuous variable was assessed using Shapiro-Wilk tests and visual inspection of Q-Q plots. Normally distributed variables were reported as mean ± standard deviation and compared within individuals using paired t-test. Non-normally distributed continuous variables were reported as medians with interquartile range and compared within individuals using Wilcoxon signed-rank test. Count data were reported as absolute numbers and percentages and compared within individuals with McNemar’s test. Differences between unvaccinated/vaccinated and individuals with/without new symptoms were analysed using unpaired t-tests, Wilcoxon rank-sum and chi-square tests as appropriate. Continuous outcomes were analysed with multiple linear regression, and at follow-up, adjusted for baseline values (ANCOVA approach). Model assumptions (residual normality, linearity, homoscedasticity and absence of multicollinearity) were evaluated using residual plots, variance inflation factors and formal tests as appropriate. For outcomes with non-normal residuals we used quantile (median) regression. Binary outcomes were modelled with logistic regression. Linearity of continuous predictors in the logit was assessed by graphical inspection, and model fit was evaluated with the Hosmer–Lemeshow statistic. Sensitivity analyses excluding the four medicated individuals at baseline and the eight unmedicated individuals at follow-up produced no overall differences i.e. they were retained in the primary analyses. Two-sided p-values below 0.05 were regarded as statistically significant, unadjusted for multiple testing because of the exploratory nature of the study design; consequently, statistically significant results should be interpreted with caution.

## Results

Demographic and laboratory characteristics are shown in Table 1. The included cohort (n=130) was predominantly of European ancestry, and all were HLA-DQB1\**06:02* positive. CSF hypocretin-1 measurements were available for 126 individuals: 83 had undetectable levels (<40 pg/ml) and 40 had low but detectable levels (40-150 pg/ml). Three individuals, reported only as “low hypocretin-1”, were excluded from hypocretin deficiency severity analyses. One hundred thirteen individuals were Pandemrix®-vaccinated. Four individuals (three vaccinated, one unvaccinated) were immunomodulated prior to/immediately after baseline (2/4 are previously reported [62]). Individuals lost to follow-up i.e. excluded; (n=23) did not differ from included individuals regarding age, sex and core NT1 features.

**Table 1:** Demographic and laboratory characteristics of post-H1N1 NT1 individuals.

| **Demographic characteristics <sup>a</sup>** | **NT1 individuals**<br>**n=130** |
| --- | --- |
| Sex (female) | 83 (63.8%) |
| Age at disease onset (years), mean ± SD | 16.8 ± 11.3 (range: 2.0 - 54.4) |
| Diagnostic delay (years), mean ± SD | 3.2 ± 2.3 (range: 0.1 – 10-3) |
| Age at baseline (years), mean ± SD | 23.0 ± 12.0 (range: 6.4 - 61.8) |
| Disease duration at baseline (years), mean ± SD | 6.2 ± 2.8 (range: 0.2 - 11.6) |
| Age at follow-up (years), mean ± SD | 28.5 ± 11.6 (range: 13.2 – 67.6) |
| Disease duration at follow-up (years), mean ± SD | 11.7 ± 2.0 (range: 4.1 – 14.3) |
| Follow-up time (years), mean ± SD | 5.5 ± 1.4 (range: 2.4 - 7.0) |
| **Laboratory characteristics** |  |
| HLA-DQB1*06:02 positive (yes), *n* (%) | 130 (100.0%) |
| Pandemrix<sup>®</sup>-vaccinated (yes), *n* (%) | 113 (86.9%) |
| CSF-Hypocretin-1 level (pg/ml)<sup>b</sup> |  |
| Low but detectable (40-150), *n* (%) | 40 (31.7%) |
| Undetectable <40, *n* (%) | 83 (65.9%) |
| Unspecified <150, *n* (%) | 3 (2.4 %) |

NT1= narcolepsy type 1. SD= standard deviation. HLA= human leukocyte antigen. CSF= cerebrospinal fluid.
<sup>a</sup> The individuals were predominantly of European ancestry, including three individuals of mixed Sámi-European ancestry, one African-European, one Arab-European, and one East Asian-European ancestry. <sup>b</sup> In four individuals CSF-hypocretin-1 level was not measured; all of these had cataplexy and were HLA-DQB1\*06:02 positive. One individual had borderline values of 150 and 152 pg/ml and was included in the low but detectable group.

### Phenotype characteristics at baseline

At baseline, mean ESS score was high (18/24) (Table 2). Cataplexy was the most prevalent symptom after EDS (lifetime occurrence of symptoms was 100% for EDS, 95 % for cataplexy, 83 % for hypnagogic hallucinations, and 78 % for sleep paralysis). Daily cataplexy was reported by 52 %, daily hypnagogic hallucinations by 20 %, and daily sleep paralysis by 6 % (Figure 1). PSG/MSLT measures showed high objective sleepiness with a mean MSLT SL of 2.05 minutes, moreover median 5/5 SOREMPs on MSLT, and fragmented night sleep (mean SSSI: 14.4 ± 4.0; mean AI: 2.4 ± 1.2) (Table 2). Overweight and obesity were common: 30 % of children were overweight and 19 % obese; 34 % of adults were overweight and 38 % were obese (Figure 2).

**Figure 1:**
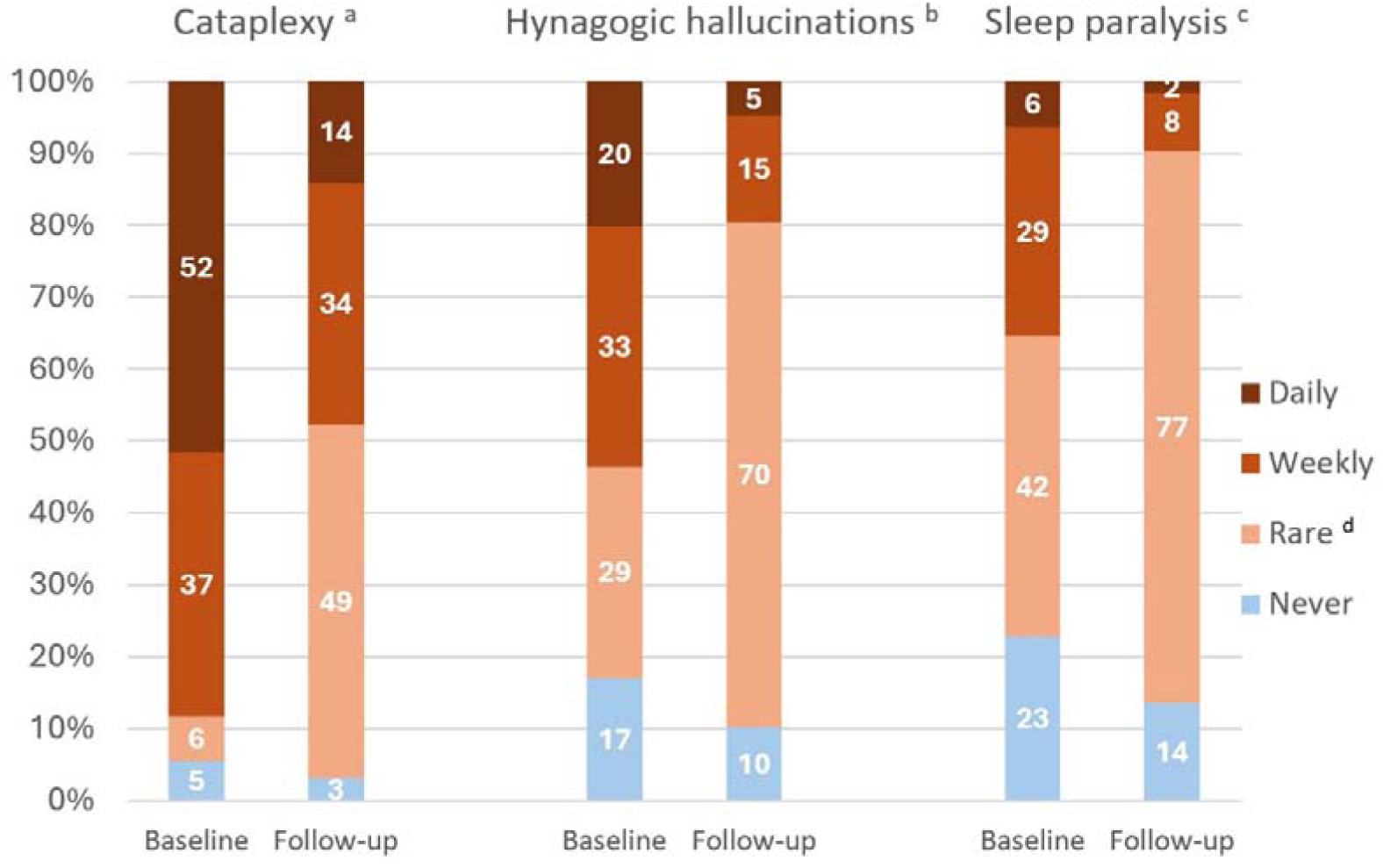
Frequency of core symptoms at baseline and follow-up ^a^ Frequency of cataplexy is missing for one individual at baseline, one individual at follow-up and one individual at both baseline and follow-up. ^b^ Frequency of hypnagogic hallucinations is missing for one individual at baseline and two individuals at follow-up. ^c^ Frequency of sleep paralysis is missing for three individuals at baseline and five individuals at follow-up. ^d^ The “rare” category includes all individuals with lifetime occurrence of the symptom and with frequency less than once per week.

**Figure 2.**
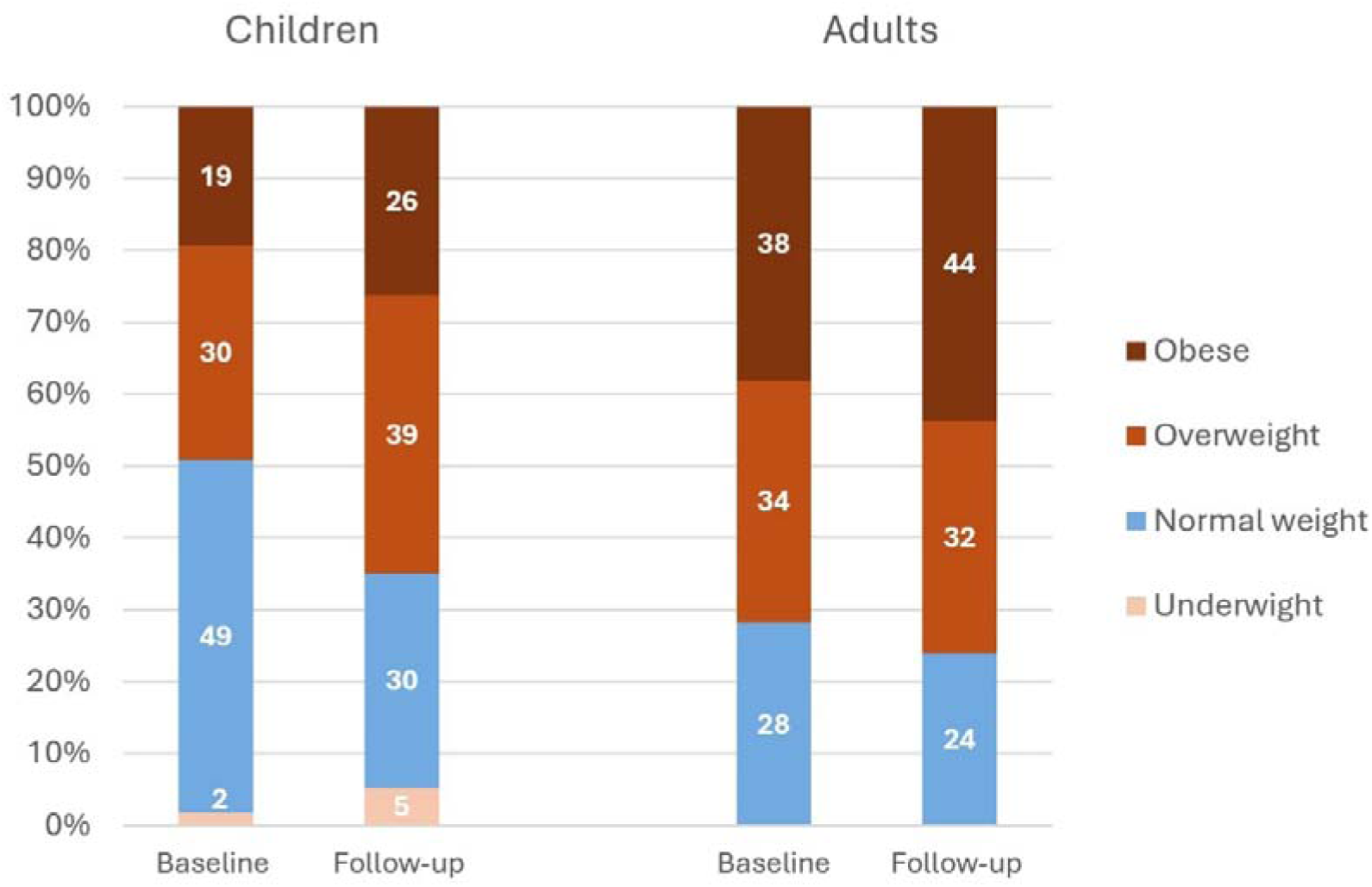
B**M**I **categories in children and adults at baseline and follow-up** NT1= narcolepsy type 1. BMI = body mass index. At both baseline and follow-up, the children group includes individuals with age < 18 years at baseline, and the adult group includes individuals with age ≥ 18 years at baseline. BMI data were missing for one of 58 children at baseline and one of 58 children at follow-up (different individuals missing data). Among adults, BMI data were missing for one of 72 adults at baseline and for two of 72 adults at follow-up (different individuals missing data). BMI categories for children are presented according to International Obesity Task Force-thresholds [59].

**Table 2:** Core symptoms, PSG/MSLT, and BMI measures at baseline and follow-up.

|  | Baseline <sup>a</sup><br>n= 130 | Follow-up<br>n= 130 | p-value |
| --- | --- | --- | --- |
| Epworth Sleepiness Scale <sup>b</sup>, mean ± SD | 18.0 ± 3.8 | 14.1 ± 4.4 | <0.001*** |
| Cataplexy (lifetime occurrence), *n* (%) | 123 (94.6 %) | 126 (96.9 %) | 0.083 |
| Hypnagogic hallucinations (lifetime occurrence), *n* (%) | 108 (83.1 %) | 117 (90.0) | 0.003** |
| Sleep paralysis (lifetime occurrence), *n* (%) | 101 (77.7 %) | 113 (86.9 %) | 0.001** |
| PSG SOREMP <sup>c</sup> (yes) | 76 (59.8%) | 59 (45.7%) | 0.007** |
| PSG SSSI <sup>c</sup>, mean ± SD | 14.4 ± 4.0 | 15.6 ± 5.0 | 0.013** |
| PSG AI <sup>c</sup>, mean ± SD | 2.4 ± 1.2 | 3.0 ± 1.7 | < 0.001*** |
| MSLT no. of SOREMPs <sup>c</sup>, median, IQR | 5 (4-5) | 2 (1-4) | < 0.001*** |
| MSLT mean SL <sup>c</sup> (min), median, IQR | 2.05 (0.9 – 3.5) | 4.6 (2.6-7.8) | < 0.001*** |
| BMI in children <sup>d</sup>, mean ± SD | 23.6 ± 4.7 | 27.3 ± 6.1 | < 0.001*** |
| BMI in adults <sup>d</sup>, mean ± SD | 28.4 ± 5.5 | 29.1 ± 5.9 | 0.095 |

SD= standard deviation. PSG = polysomnography. SOREMP(s) = sleep onset rapid eye movement period(s). SSSI= sleep stage shift-index. AI= awakening-index. MSLT= multiple sleep latency test. IQR= interquartile range. SL= sleep latency. BMI = body mass index.
\* $p$ -values $\leq 0.05$ . \*\* $p$ -values $\leq 0.01$ . \*\*\* $p$ -values $\leq 0.001$ .
<sup>a</sup> Four individuals did not discontinue medication at baseline. <sup>b</sup> One individual lacked the ESS score at follow-up. <sup>c</sup> PSG and MSLT recordings were unavailable at baseline for three individuals: two due to technically invalid recordings and one because the individual declined to complete the sleep recordings. At follow-up, PSG and MSLT recordings were missing for another individual who declined to complete the measurements. <sup>d</sup> At both baseline and follow-up, the children category includes individuals with age < 18 years at baseline, and the adult category includes individuals with age $\geq 18$ years at baseline. BMI data were missing for one of 58 children at baseline and one of 58 children at follow-up (different individuals missing data). Among adults, BMI data were missing for one of 72 adults at baseline and for two of 72 adults at follow-up (different individuals missing data).

### Factors associated with phenotype severity at baseline

Factors associated with baseline phenotype severity are summarized in Tables 3 and 4. Female sex was associated with greater symptom burden: women had higher odds of daily cataplexy (OR=4.24, p=0.001 Table S1) and trends for higher ESS scores (β= 1.94, *p*=0.066, Table 3) and shorter mean sleep latency (β=-0.97, *p*=0.073, Table 3) than males. Younger age was associated with higher objective sleepiness (mean MSLT SL increased with age (β=0.06, *p*=0.017, Table 4)), predicted more MSLT SOREMPs (β=-0.03, *p*=0.001, Table 4), and showed a trend for having a PSG SOREMP (OR=1.97, *p*=0.073 Table 4). Disease duration was not associated with symptoms or PSG/MSLT measures.

**Table 3:**
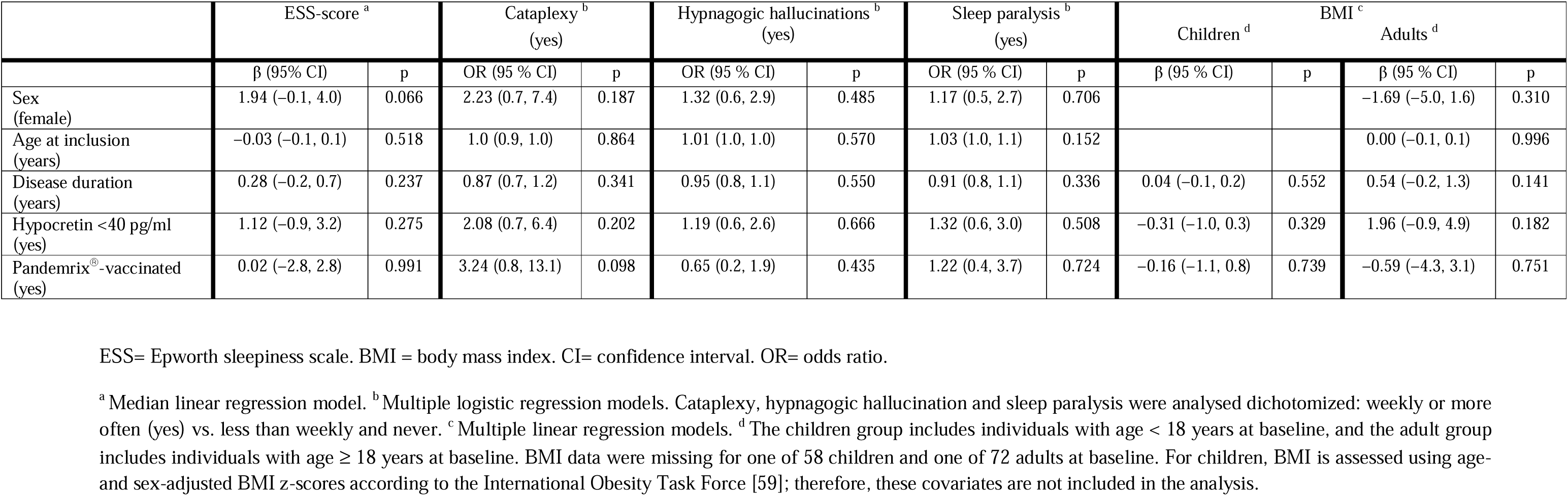
Predictors for symptom and BMI severity at baseline.

**Table 4:** Predictors for PSG and MSLT severity at baseline.

|  | PSG SSSI <sup>a</sup> |  | PSG AI <sup>a</sup> |  | PSG SOREMP <sup>b</sup> (yes) |  | Mean MSLT SL <sup>c</sup> (min) |  | No. of MSLT SOREMPs <sup>a</sup> |  |
| --- | --- | --- | --- | --- | --- | --- | --- | --- | --- | --- |
| | $\beta$ (95 % CI) | p | $\beta$ (95 % CI) | p | OR (95 % CI) | p | $\beta$ (95 % CI) | p | $\beta$ | p |
| Sex (female) | -0.77 (-2.4, 0.9) | 0.357 | 0.04 (-0.4, 0.5) | 0.849 | 1.71 (0.7, 4.0) | 0.221 | -0.97 (-2.0, 0.1) | 0.073 | -0.05 (-0.4, 0.3) | 0.792 |
| Age at inclusion (years) | 0.04 (0.0, 0.1) | 0.334 | 0.02 (0.0, 0.0) | 0.107 | 1.97 (0.9, 1.0) | 0.073 | 0.06 (0.0, 0.1) | 0.017* | - 0.03 (-0.1, 0.0) | 0.001**<br>* |
| Disease duration (years) | -0.1 (-0.5, 0.3) | 0.599 | 0.00 (-0.1, 0.1) | 0.938 | 0.92 (0.8, 1.1) | 0.400 | 0.01 (-0.2, 0.3) | 0.912 | 0.05 (-0.0, 0.1) | 0.220 |
| Hypocretin <40 pg/ml (yes) | -0.38 (-2.0, 1.2) | 0.644 | 0.23 (-0.2, 0.7) | 0.301 | 2.38 (1.0, 5.5)* | 0.04* | -1.13 (-2.2, -0.1) | 0.034* | 0.21 (-0.2, 0.6) | 0.266 |
| Pandemrix<sup>®</sup> -vaccinated (yes) | 0.6 (-1.6, 2.8) | 0.586 | 0.56 (-0.0, 1.2) | 0.061 | 2.93 (1.0, 9.0) | 0.061 | -1.43 (-2.9, -0.0) | 0.049* | 0.32 (-0.2, 0.8) | 0.216 |

PSG = polysomnography. SSSI= sleep stage shift index. AI= awakening index. SOREMP(s) = sleep onset rapid eye movement period(s). MSLT= multiple sleep latency test. SL= sleep latency. CI= confidence interval. OR= odds ratio.
\* $p$ -values $\leq 0.05$ . \*\* $p$ -values $\leq 0.01$ . \*\*\* $p$ -values $\leq 0.001$ .
<sup>a</sup> Multiple linear regression models. <sup>b</sup> Multiple logistic regression model. <sup>c</sup> Median linear regression model.

Hypocretin deficiency severity was not associated with any symptom severity but was associated with more severe objective measures: individuals with undetectable hypocretin (< 40 pg/ml) had shorter mean MSLT SL (β= - 1.13, *p*=0.034, Table 4) and higher odds of a PSG SOREMP (OR=2.38, p=0.04, Table 4) than those with low but detectable hypocretin levels (40–150 pg/ml). Individuals with a higher BMI were more often male (β =-3.41, p=0.024, Table S2) and had significantly higher ESS score (adults: β = 0.50, p = 0.002; children: β = 0.09, p=0.033, Table S2).

### Phenotype course from baseline to follow-up

When analysing differences from baseline to follow-up there was an overall improvement in both symptoms and PSG/MSLT measures (Table 2). Mean ESS scores decreased from 18/24 to approximately 14/24 (*p* < 0.001). Daytime sleepiness also objectively improved as median MSLT SL increased from 2.1 to 4.6 minutes (p<0.001), moreover median number of MSLT SOREMPs decreased from 5 to 2 (p<0.001). The proportion of individuals with a PSG SOREMP decreased from 60 % at baseline to 46 % at follow-up (p=0.007). At the group level, symptom severity (frequency) decreased for cataplexy, hypnagogic hallucinations and sleep paralysis (Figure 1). In contrast, PSG sleep fragmentation slightly worsened: SSSI increased from 14.4 to 15.6 (*p* = 0.013) and AI increased from 2.4 to 3.0 (*p* < 0.001). A small subgroup had developed new symptoms during follow-up (three developed cataplexy, nine developed hypnagogic hallucinations; thirteen developed sleep paralysis); these individuals had significantly shorter median disease duration at baseline (5.5 (2.5–6.9) vs. 6.3 (5.2-7.7) years in the remaining group, *p=*0.041 (Table S3).

The proportion of overweight in the children group (i.e. individuals <18 years at baseline) increased from 30% at baseline to 39 % at follow-up, and obesity increased from 19 % to 26 % (Figure 2). Mean BMI in the children group increased from 23.6 to 27.3 (*p*<0.001) (Table 2). In adults (≥18 years at baseline), overweight percentage remained stable at 34 % at baseline and 32 % at follow-up, obesity increased from 38 % to 44 %, and mean BMI did not significantly change (but tended to increase): 28.4 at baseline vs. 29.1 at follow-up; *p* = 0.095 (Table 2).

### Factors associated with phenotype severity at follow-up

Table 5 and 6 summarize factors associated with phenotype severity at follow-up. For all PSG/MSLT measures and all symptoms (except hypnagogic hallucinations) the corresponding baseline value strongly predicted the same outcome at follow-up. Female sex remained associated with higher ESS score at follow-up (β = 2.01, *p*=0.012) (Table 5) but no longer associated with daily cataplexy (Table S1). Older age was associated with increased sleep fragmentation as measured by SSSI (β = 0.14 per year, p=0.001, Table 6). The baseline associations with younger individuals having shorter mean MSLT SL and more SOREMPs were attenuated at follow-up (associations with mean MSLT SL no longer significant; borderline associated with more MSLT SOREMPs (β =-0.03, *p*=0.099, Table 6)). Disease duration was not associated with follow-up outcomes apart from a borderline association with higher SSSI (β = 0.42, *p* = 0.068, Table 6). Mean MSLT SL was shorter in adults with higher BMI (β =-0.28, p = 0.049, Table S2; not found in children). The association between the most severe hypocretin deficiency (<40 pg/ml) and shorter mean MSLT SL) found at baseline did not persist at follow-up (β =-1.59, *p* = 0.164, Table 6), but was borderline associated with a higher odds ratio of more frequent hypnagogic hallucinations at follow-up (OR=3.37, *p*=0.073, Table 6).

**Table 5:**
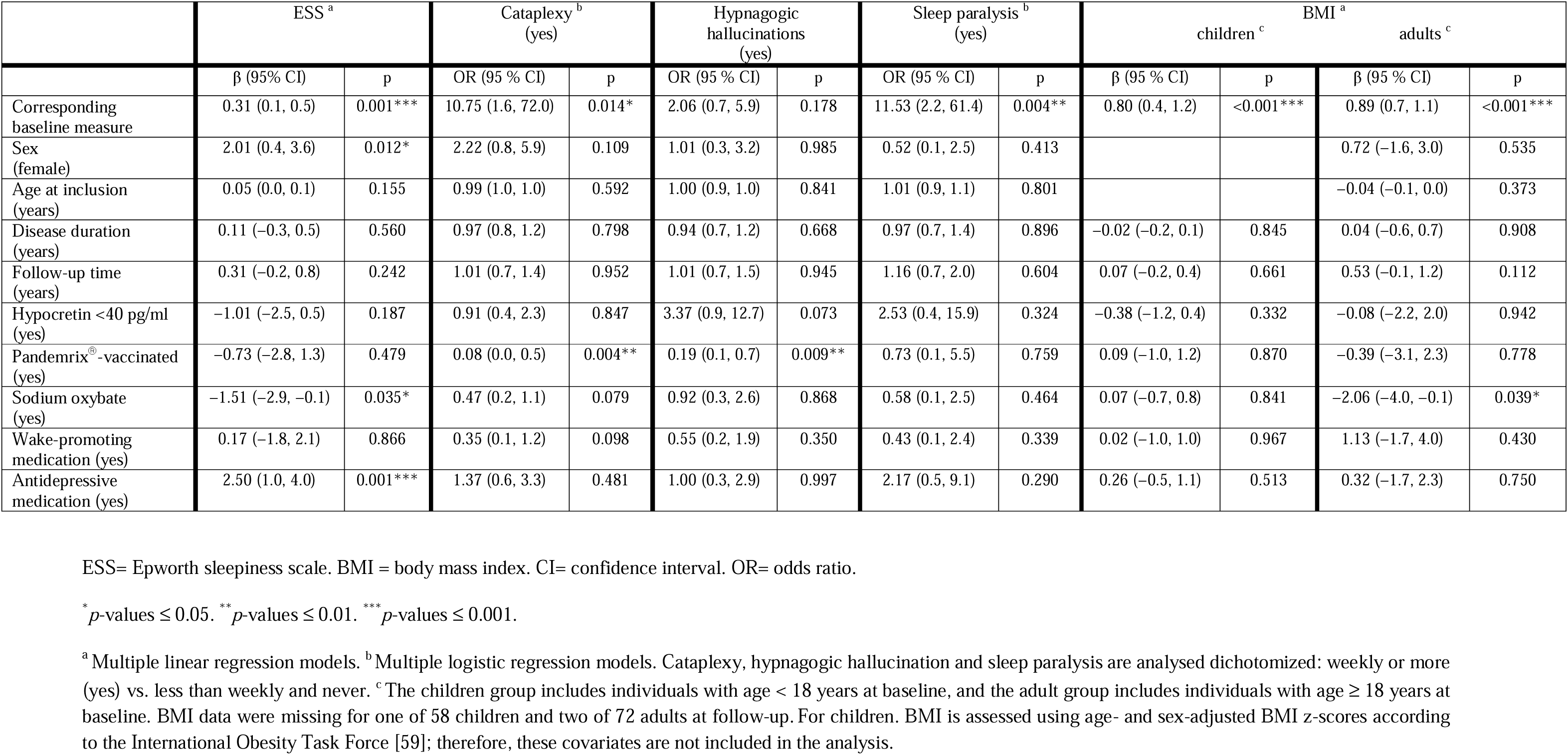
Predictors for symptom and BMI severity at follow-up.

**Table 6:**
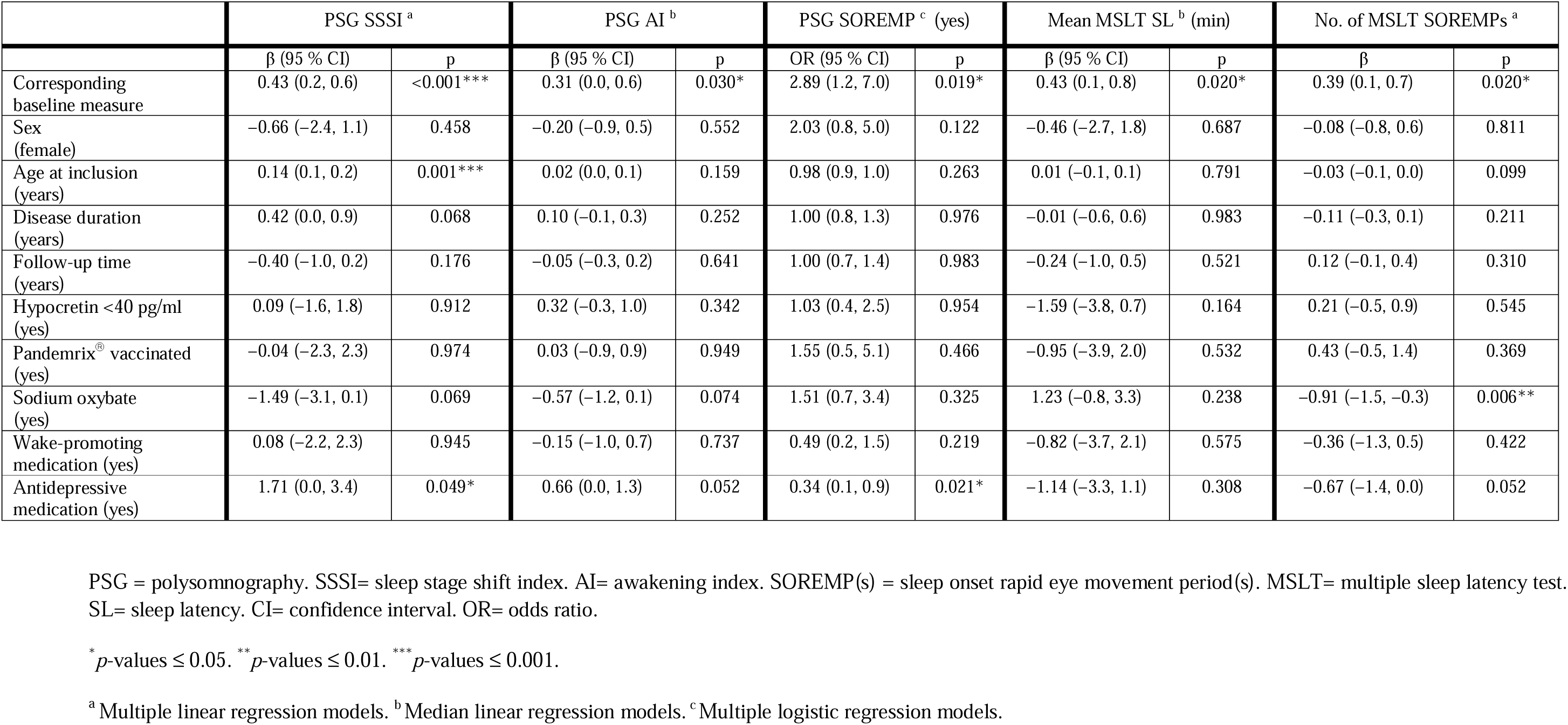
Predictors for PSG and MSLT severity at follow-up.

Regarding medication at follow-up, 99 individuals were on combination therapy, 23 on monotherapy and 8 were unmedicated (Table 7). Multivariate regression analyses including medication are shown in Tables 5 and 6.

**Table 7:** Medication used at follow-up.

| Medication <sup>a</sup> | n (%) |
| --- | --- |
| Sodium oxybate | 63 (48.5%) |
| Pitolisant | 21 (16.2%) |
| Modafinil | 40 (30.8%) |
| Methylphenidate | 75 (57.7%) |
| Other central stimulants <sup>b</sup> | 13 (10.0%) |
| SNRI | 36 (27.7%) |
| Other antidepressive medication <sup>c</sup> | 9 (6.9%) |
| Antipsychotic medication | 2 (1.5%) |
| Hypnotic medication | 3 (2.3%) |
| Melatonin | 4 (3.1%) |
| No medication <sup>c</sup> | 8 (6.2 %) |

SNRI = Serotonin–norepinephrine reuptake inhibitor
<sup>a</sup> Ninety-nine individuals were on multiple medications. <sup>b</sup> Dexamphetamine, Lisdexsamphetamine, Solriamfetol. <sup>c</sup> Selective serotonin reuptake inhibitor, tricyclic antidepressant, dopamine–norepinephrine reuptake inhibitor. <sup>c</sup> Two women in their first trimester of pregnancy had temporarily stopped medication.

### Pandemrix®-vaccinated versus unvaccinated NT1

Demographic, laboratory, and medication characteristics +/- Pandemrix®-vaccination status are shown in Table S4. Unvaccinated individuals had significantly shorter disease duration at baseline and follow-up. Vaccinated individuals were numerically (but not significantly) younger at disease onset (median 12.7 years (9.9 – 16.6)) than unvaccinated NT1 individuals (median 15.4 years (12.5 – 25.6)) and showed a trend for more likely being medicated at follow-up (p=0.069).

Phenotype features +/- vaccination status are shown in Table 8 and illustrated in Figures S1-S4. At baseline, vaccinated individuals had numerically (though not significantly/borderline significantly) more severe PSG/MSLT measures. In the baseline multivariate models, vaccination was significantly associated with having shorter mean MSLT SL (β=-1.43, *p*=0.049) and tended to have higher odds for a PSG SOREMP (β=2.93, *p*=0.061) and higher AI (β=0.56, *p*=0.061) (Table 4). Vaccinated individuals also overall had numerically (but not significantly) more symptoms (higher ESS and lifetime occurrence of cataplexy and hypnagogic hallucinations, Table 8), and in the multivariate regression models vaccination was borderline associated with more frequent cataplexy (OR=3.24, p=0.098, Table 3).

**Table 8:** Symptoms and PSG/MSLT measures in H1N1 (Pandemrix®)-vaccinated versus unvaccinated NT1 individuals at baseline and follow-up.

|  | Baseline |  |  | Follow-up |  |  |
| --- | --- | --- | --- | --- | --- | --- |
|  | Pandemrix®-vaccinated<br>n= 113 | Unvaccinated<br>n= 17 | p | Pandemrix®-vaccinated<br>n= 113 | Unvaccinated<br>n= 17 | p |
| Epworth Sleepiness Scale (ESS), mean ± SD | 18.1 ± 3.9 | 17.5 ± 3.1 | 0.518 | 14.0 ± 4.6<sup>a</sup> | 14.7 ± 3.4 | 0.604 |
| Cataplexy (lifetime occurrence), *n* (%) | 108 (95.6 %) | 15 (88.2) | 0.228 | 110 (97.3 %) | 16 (94.1 %) | 0.443 |
| Hypnagogic hallucinations (lifetime occurrence), *n* (%) | 96 (85.0 %) | 12 (70.6 %) | 0.165 | 102 (90.2 %) | 15 (88.2 %) | 0.679 |
| Sleep paralysis (lifetime occurrence), *n* (%) | 88 (77.9) | 13 (76.5 %) | 1.000 | 97 (85.8 %) | 16 (94.1 %) | 0.698 |
| Weekly cataplexy<sup>b</sup> (yes), *n* (%) | 100 (90.1 %) <sup>c</sup> | 13 (76.5 %) | 0.115 | 47 (42.3 %) <sup>c</sup> | 14 (82.4 %) | 0.003** |
| Weekly hypnagogic hallucinations<sup>b</sup> (yes), *n* (%) | 58 (51.8 %) <sup>d</sup> | 11 (64.7) | 0.320 | 17 (15.3 %) <sup>d</sup> | 8 (47.1 %) | 0.002** |
| Weekly sleep paralysis<sup>b</sup> (yes), *n* (%) | 39 (35.5 %) <sup>e</sup> | 6 (35.3 %) | 0.990 | 10 (9.1 %) <sup>e</sup> | 2 (12.5 %) | 0.673 |
| PSG SOREMP (yes), *n* (%) | 69 (62.7%) <sup>f</sup> | 7 (41.2 %) | 0.092 | 53 (47.3%) <sup>f</sup> | 6 (35.3 %) | 0.354 |
| PSG SSSI, mean ± SD | 14.5 ± 3.9 <sup>f</sup> | 13.9 ± 4.9 | 0.575 | 15.6 ± 5.0 <sup>f</sup> | 15.2 ± 5.2 | 0.718 |
| PSG AI, mean ± SD | 2.5 ± 1.3 <sup>f</sup> | 1.9 ± 1.3 | 0.063 | 3.0 ± 1.7 <sup>f</sup> | 2.6 ± 1.5 | 0.375 |
| MSLT no. of SOREMPs, median (IQR) | 5 (4-5) <sup>g</sup> | 5 (4-5) | 0.361 | 3 (0-4) <sup>g</sup> | 2 (1-4) | 0.629 |
| mean ± SD | 4.5 ± 0.9 <sup>g</sup> | 4.1 ± 1.4 |  | 2.6 ± 1.8 <sup>g</sup> | 2.4 ± 2.2 |  |
| MSLT mean SL (min), median (IQR) | 1.9 (0.8-3.3) <sup>g</sup> | 2.4 (1.8-5.7) | 0.089 | 4.5 (2.6-7.7) <sup>g</sup> | 5.3 (4.1 – 9.2) | 0.305 |
| BMI in children<sup>h</sup>, mean ± SD | 23.7 ± 4.8 | 22.9 ± 4.2 | 0.70 | 27.5 ± 5.9 | 25.6 ± 7.6 | 0.478 |
| BMI in adults<sup>h</sup>, mean ± SD | 28.3 ± 5.7 | 27.5 ± 4.2 | 0.653 | 29.0 ± 6.1 | 29.3 ± 4.6 | 0.912 |

SD= standard deviation. PSG = polysomnography. SOREMP(s) = sleep onset rapid eye movement period(s). SSSI= sleep stage shift index. AI= awakening index. MSLT= multiple sleep latency test. IQR= interquartile range. SL= sleep latency. BMI = body mass index.
\* $p$ -values $\leq 0.05$ . \*\* $p$ -values $\leq 0.01$ . \*\*\* $p$ -values $\leq 0.001$ .

At follow-up, lifetime occurrence of symptoms was overall similar in vaccinated and unvaccinated individuals (Table 8), but the two groups showed different longitudinal patterns in symptom frequency, as vaccinated individuals had greater improvement in cataplexy and hypnagogic hallucinations frequency at follow-up (lower odds of weekly cataplexy (OR = 0.08, p = 0.004) and hypnagogic hallucinations (OR = 0.19, p = 0.009) in the multivariate models (Table 5); illustrated in Figures S1-S2). Vaccination was not associated with ESS scores or sleep paralysis frequency at follow-up (Table 5; Figure S3). PSG/MSLT measures were overall still numerically worse in vaccinated individuals at follow-up (Table 8), but vaccination no longer independently predicted more severe PSG/MSLT findings in the multivariate regression model (Table 6).

## Discussion

In this large prospective cohort of 130 post-H1N1 (mainly H1N1(Pandemrix®)-vaccinated) NT1 individuals followed-up after a mean of 5.5 years, we confirm that post-H1N1 NT1 in several ways shows typical NT1 features similar to previously described in pre-H1N1/sporadic NT1, as individuals are HLA-DQB1*0602-positive, hypocretin deficient, have cataplexy and PSG/MSLT SOREMPs. Likewise, we confirm that female sex and younger age are associated with more severe phenotype features also in post-H1N1 NT1. We novelly show that severe hypocretin deficiency is associated with the presence of nighttime REM-sleep instability (PSG SOREMPs) and confirm an association with severe objective daytime sleepiness (lower MSLT SL). For the whole cohort, symptoms and PSG/MSLT phenotype features generally improve at follow-up, mainly predicted by the baseline severity of the same outcomes. Several vaccination-associated phenotype features are also found, as being vaccinated independently predicts more severe PSG/MSLT measures and borderline predicts more severe cataplexy at baseline. However, vaccinated individuals show a different long-term clinical course than unvaccinated NT1 individuals, as vaccination is independently associated with greater improvement of symptom severity/frequency and no longer associated with PSG/MSLT severity at follow-up.

Our finding of long-term improvement of PSG/MSLT measures in a post-H1N1 NT1 (mainly Pandemrix®-vaccinated) cohort is novel. This contrasts with stable PSG/MSLT measures in two small/moderate sized longitudinal sporadic NT1 cohorts, though notably these individuals were persistently unmedicated/drug naïve and in one of the studies the individuals were included closer to disease onset with a shorter follow-up [63] and the other with unspecified disease duration [43].

Likewise, that PSG sleep fragmentation worsens with increasing age is a novel finding in post-H1N1 NT1 and in concordance with sporadic NT1 studies [33,41,44]. Because sleep fragmentation is known to increase with age in the general population, it is unclear whether this reflects disease specific progression or normative aging [64,65]. As both subjective and objective measures improve (decreased ESS, less frequent cataplexy, hypnagogic hallucinations and sleep paralysis, fewer PSG/MSLT SOREMPs, increased MSLT SL) this supports a genuine phenotype severity change rather than merely altered symptom reporting. Our findings are further strengthened by the consistent testing conditions (same sleep-laboratory, PSG/MSLT equipment, staff, sleep scoring somnologist/technician). We further found that BMI is high but remains largely stable at the group level, though adolescents tend to develop overweight or obesity during transition to adulthood. Moreover, adult men have initial, but not persistent, higher BMI. This pattern mirrors the literature on sporadic NT1: some report weight gain and continued BMI-increases over time [41,43,54], while others report stabilisation of BMI with prolonged disease [45]. Our BMI findings support that clinicians should monitor adolescents and young adults with NT1 closely for weight gain during the transition to adulthood and consider structured weight-management and metabolic screening as part of long-term care.

Previous reports of Pandemrix®-vaccinated NT1 individuals with early and fulminant disease onset raised the possibility of a distinct, more severe vaccinated subvariant [20,50]. Prior comparative studies have inconsistent findings. In our present cohort, the largest longitudinal post-H1N1 NT1 study to date, we confirm that vaccinated individuals have significantly more severe objective sleepiness at baseline (shorter MSLT SL) and trends for more MSLT SOREMPs, more fragmented sleep, and for more severe cataplexy. However, the phenotype of vaccinated individuals also improves more over time: greater symptom improvement (lower odds of frequent cataplexy and hypnagogic hallucinations), and – although PSG/MSLT measures remain numerically more severe than in unvaccinated individuals - vaccination no longer independently predicts PSG/MSLT severity at follow-up. Our cohort has high data completeness/availability of the known NT1 phenotype biological predictors (HLA-typing in 100%; CSF hypocretin-1 measures in 97%). The different clinical course in vaccinated and unvaccinated NT1 is not explained by HLA (all individuals are HLA-DQB1*06:02-positive) or by major differences in hypocretin deficiency severity (undetectable hypocretin levels in 70 % vaccinated vs. 64.7% in unvaccinated individuals (χ^2^(1) = 0.185, *p*=0.667). As PSG/MSLT measures are associated with vaccination status at baseline but no longer at follow-up, one hypothesis may be that Pandemrix®-vaccination initially triggers a relatively intense acute immune-mediated hypocretin-producing neuron loss/hypocretin system dysregulation, producing a more severe early phenotype that over time partially undergoes compensatory adaptation. Our previous MRI finding of more widespread Diffusion Tensor Imaging (DTI) changes (in a subset of the present baseline cohort, n= 57) [66] than in studies of sporadic NT1 (n= 12; n=22, respectively) [67] [68] could support this. However, as the most predictive factor for long-term phenotype severity is the same outcomes at baseline – and as vaccination predicts the baseline outcomes - it is also possible that vaccination indirectly still play a long-term role. Alternative explanations include differences in healthcare seeking behaviour, psychosocial factors, and medication (e.g. the trend for higher %-medicated vaccinated individuals) which cannot be excluded by our observational data.

It is well-known that hypocretin deficiency is typical for NT1. However – because hypocretin-1 can only be measured in CSF and is (though recommendable) still not obligatory for NT1 diagnostics - many clinical NT1 cohorts have limited/incomplete CSF hypocretin-1 data. Whether the severity of hypocretin deficiency (undetectable versus low but detectable CSF hypocretin-1 levels) predicts a long-term more severe NT1 phenotype in humans is therefore largely unknown. We here show that undetectable hypocretin levels are associated with nightly REM-instability (PSG SOREMP) and worse objective daytime sleepiness (lower mean MSLT SL) at baseline but not associated with phenotype severity at follow-up in post-H1N1 NT1. To our knowledge this has not been investigated longitudinally in sporadic nor in post-H1N1 NT1, but undetectable hypocretin levels are in a cross-sectional cohort study associated with low MSLT SL and MSLT SOREMPs in partial accordance with our findings [37]. Our finding of association with undetectable hypocretin and PSG SOREMPs is novel, though a trend was found in a sporadic NT1 cohort [69]. Notably, hypocretin deficiency severity is not associated with subjective daytime sleepiness or any other core symptoms in our study, which is in discordance with a small sporadic NT1 cohort (n=26) [42] and the vaccinated group (n=25) of the previous longitudinal post-H1N1 study by Sarkanen et al [54]. Direct comparisons between studies are complicated by some differences in hypocretin detection limits (<20-<60 pg/ml) [37–40,42,69]. However, collectively this overall still reinforces CSF hypocretin-1’s central role in sleep–wake dysregulation which may clinically help stratify individuals for intensity of early interventions, such as more proactive treatment strategies. Our hypocretin findings are strengthened by larger sample size, high hypocretin measurement completeness, and all samples analysed by the same laboratory/protocol [61].

As females in our cohort have significantly more severe cataplexy and a trend for more severe ESS and shorter mean MSLT SL at baseline, this corresponds to similar findings in a sporadic NT1 cohort and animal studies [28,70,71]. That women also have long-term increased sleepiness (increased ESS) at follow-up is to our knowledge a novel finding. Collectively, this points to likely biological sex–based mechanisms and supports that women with NT1 may need targeted therapeutic strategies and additional psychosocial support. That younger age at disease onset predicts more severe baseline PSG/MSLT measures (shorter MSLT SL, more MSLT SOREMPs, a trend for PSG SOREMPs), although these associations are attenuated at follow-up, is consistent with findings in sporadic NT1 cohorts [31–34].

Lifetime occurrence of cataplexy, hypnagogic hallucinations and sleep paralysis increased modestly over follow-up due to a subgroup with evolving symptoms. These individuals had a significantly shorter disease duration at baseline (median 5.5 years) compared to the remaining group (median 6.3 years), in overall concordance with sporadic NT1 where the majority of individuals develop their final symptom set within 5 years after onset [72].

Important limitations must be acknowledged. Follow-up was performed while individuals were on usual treatment and baseline was largely drug-free. This observational design complicates separation of natural history from treatment effects i.e. medication associations should only be regarded as hypothesis generating. However, while it is likely to assume that medication plays a role in phenotype improvement at follow-up in our study, baseline (unmedicated) subjective and objective measures strongly predicting the same follow-up (medicated) measures, and development of new symptoms between baseline and follow-up in some individuals (95% were medicated) points to that natural history is still reflected. CSF hypocretin-1 was measured at variable intervals relative to disease onset/baseline inclusion, limiting precision for longitudinal interpretation. Ceiling and floor effects in symptoms and PSG/MSLT severity measures (e.g., at baseline, approx. 88 % had frequent (at least weekly) cataplexy, 4–5/5 SOREMPs and/or low MSLT SL) may limit some association detections. Likewise, a possible floor effect due to the high prevalence of individuals with undetectable hypocretin levels may have limited possible detection of dose-response relationships. Subgroup comparisons, particularly with the relatively small unvaccinated NT1 group, have limited power i.e. increased risk of type 2 error.

In conclusion, we demonstrate that although post-H1N1 NT1 shows several phenotypical similarities to sporadic NT1 (including severity predictive factors like female sex and age), the phenotype severity and long-term clinical course are different in H1N1(Pandemrix®)-vaccinated individuals. Vaccinated individuals initially have more severe subjective and objective phenotype measures, but these improve more than in sporadic individuals. Consequently, over time vaccination no longer independently predicts phenotype severity. However, as long-term phenotype severity is most strongly predicted by the baseline phenotype severity, this may suggest possible indirect vaccination long-term effects, though medication effects cannot be excluded. We further show that severe hypocretin deficiency predicts more nightly REM-instability and higher daytime sleepiness. Our findings underscore heterogeneity in NT1 disease trajectories and the need to consider subgroups in diagnosis, prognosis, management, and research.

## Data Availability

All data produced in the present study are available upon reasonable request to the authors

## Acknowledgement

We especially thank all the individuals with NT1 that have participated in the study. We thank Ranveig Østrem for biobanking and analyses. The study was funded by a grant from the South-Eastern Norway Regional Health Authority (R.V. grant: 2017070; H.T.J. grant: 2019032) and the Norwegian Ministry of Health and Care Services (K.L, L.F.F, R.V., J.V., H.T.J., S.K.H.).

## Data Availability Statement

The data used in this article cannot be shared publicly since we do not have ethical approval to share the data due to the privacy of study participants.

## List of Abbreviations

AASM: American Academy of Sleep Medicine
AI: Awakening Index
BMI: Body Mass Index
CNS: Central Nervous System
CSF: Cerebrospinal Fluid
DTI: Diffusion Tensor Imaging
EDS: Excessive Daytime Sleepiness
ESS: Epworth Sleepiness Scale
ESRS: European Sleep Research Society
H1N1: Influenza A (H1N1) virus
HLA: Human Leucocyte Antigen
ICSD: International Classification of Sleep Disorders
ICSD-3-TR: International Classification of Sleep Disorders, 3rd Edition, Text Revision
MSLT: Multiple Sleep Latency Test
MRI: Magnetic Resonance Imaging
NT1: Narcolepsy Type 1
OR: Odds Ratio
PSG: Polysomnography
RBD: REM Sleep Behaviour Disorder
REK: Regional Committees for Medical and Health Research Ethics (Norway)
SD: Standard Deviation
SL: Sleep Latency
SSSI: Sleep Stage Shift Index
SOREMP(s): Sleep-Onset Rapid Eye Movement Period(s)
SYSVAK: Norwegian Immunization Registry
WHO: World Health Organization

