## Supplemental Figure 5 for "Course and severity of post-H1N1 narcolepsy type 1: a long-term prospective cohort study"

**Figure S5 a-b: BMI in children and adult Pandemrix®-vaccinated versus unvaccinated NT1 at baseline and follow-up**

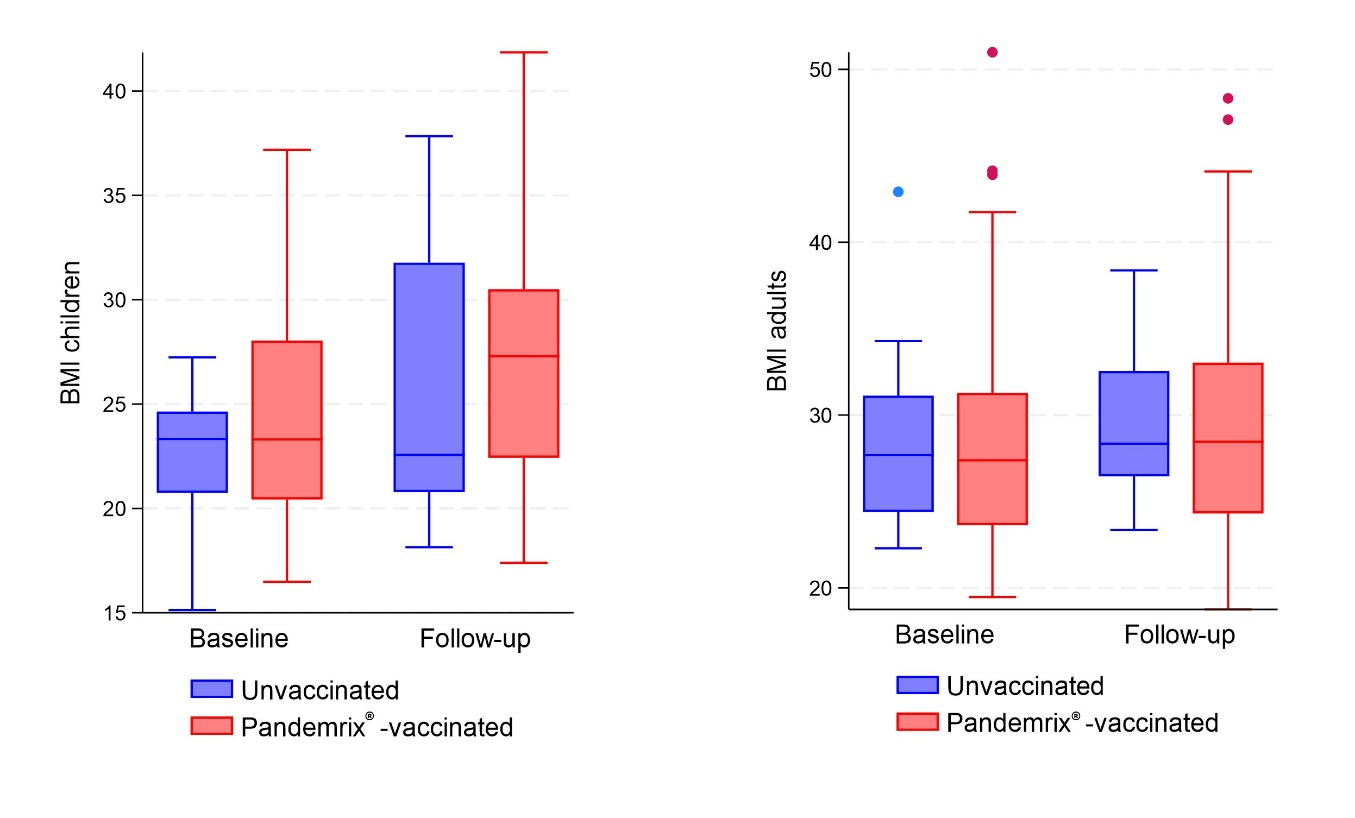

NT1= narcolepsy type 1. BMI= body mass index.

^*^*p*-values ≤ 0.05. ^**^*p*-values ≤ 0.01. ^***^*p*-values ≤ 0.001.

At both baseline and follow-up, the children group includes individuals with age < 18 years at baseline, and the adult group includes individuals with age ≥ 18 years at baseline. BMI data were missing for one of 58 children at baseline and one of 58 children at follow-up (different individuals missing data). Among adults, BMI data were missing for one of 72 adults at baseline and for two of 72 adults at follow-up (different individuals missing data).
