## Supplemental Figure 4 for "Course and severity of post-H1N1 narcolepsy type 1: a long-term prospective cohort study"

**Figure S4a-c: PSG/MSLT-measures in Pandemrix^®^-vaccinated versus unvaccinated NT1 individuals at baseline and follow-up**

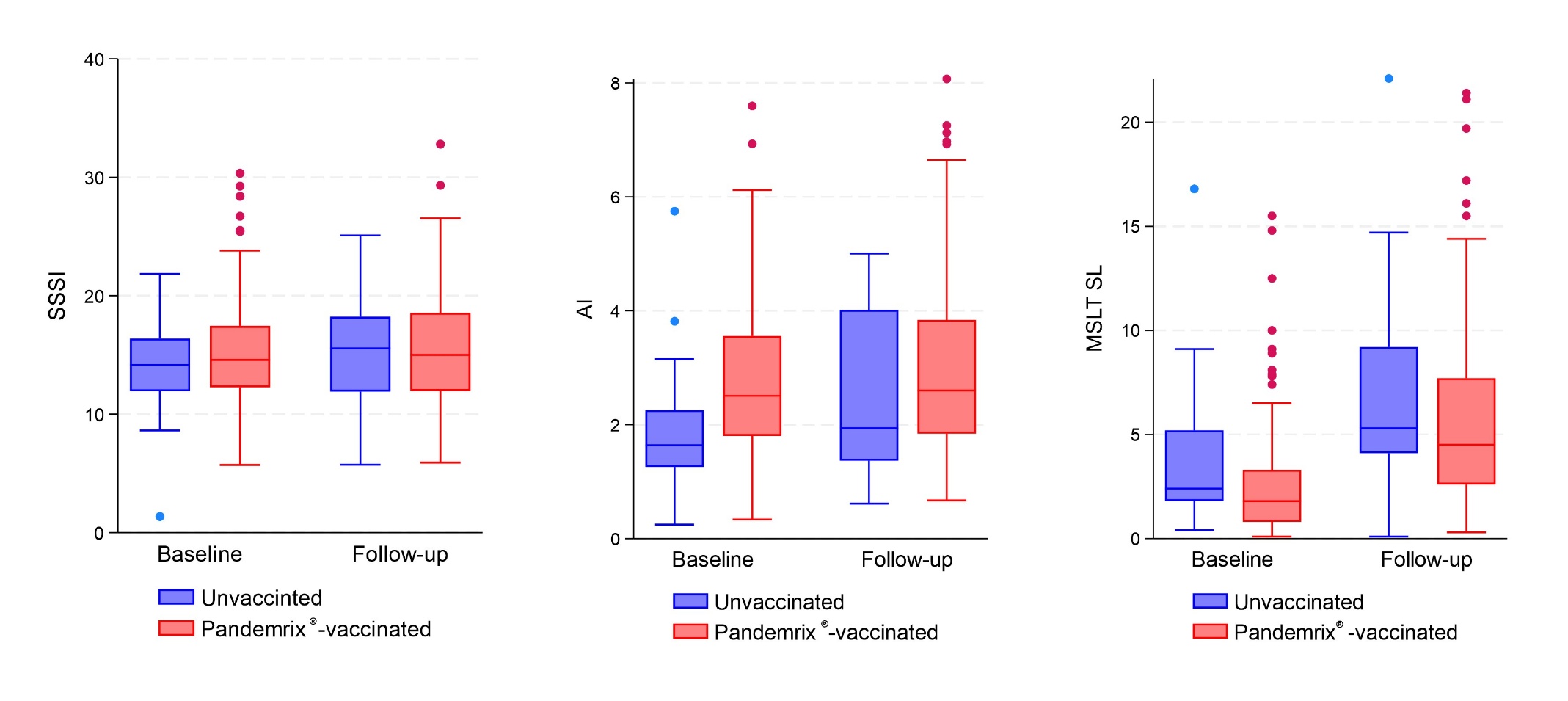

NT1= narcolepsy type 1. PSG = polysomnography. MSLT= multiple sleep latency test.SSSI= sleep stage shift index. AI= awakening index. SL= sleep latency.

^*^*p*-values ≤ 0.05. ^**^*p*-values ≤ 0.01. ^***^*p*-values ≤ 0.001.
