## Supplemental Figure 3 for "Course and severity of post-H1N1 narcolepsy type 1: a long-term prospective cohort study"

**Figure S3: ESS-scores in Pandemrix^®^-vaccinated versus unvaccinated NT1 individuals at baseline and follow-up**

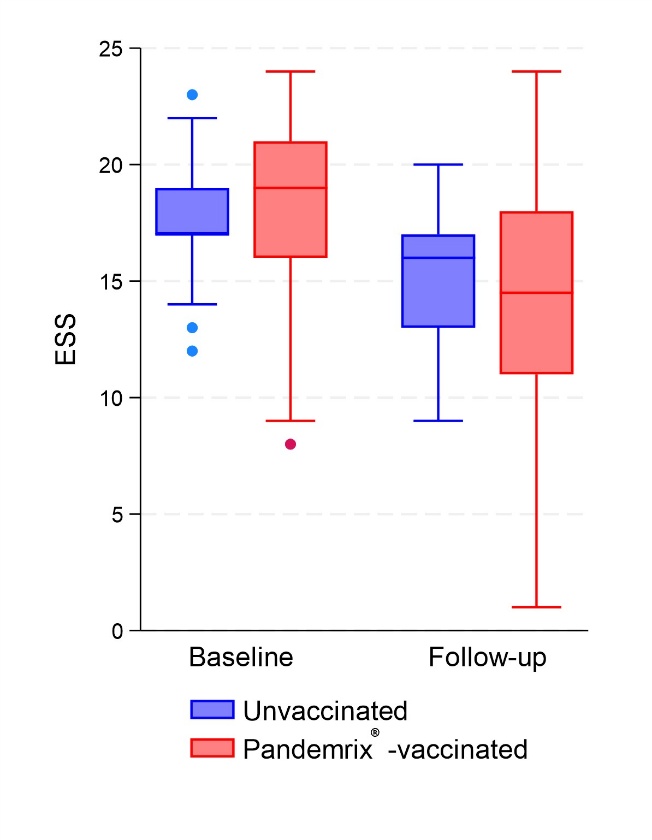

NT1= narcolepsy type 1. ESS= Epworth sleepiness scale. PSG = polysomnography. SSSI= sleep stage shift index. AI= awakening index. MSLT= multiple sleep latency test. SL= sleep latency. SOREMPs = sleep onset rapid eye movement sleep.

^*^*p*-values ≤ 0.05. ^**^*p*-values ≤ 0.01. ^***^*p*-values ≤ 0.001.
