## Supplemental Figure 2 for "Course and severity of post-H1N1 narcolepsy type 1: a long-term prospective cohort study"

**Figure S2: Daily cataplexy in Pandemrix®-vaccinated and unvaccinated NT1 individuals at baseline and follow-up**

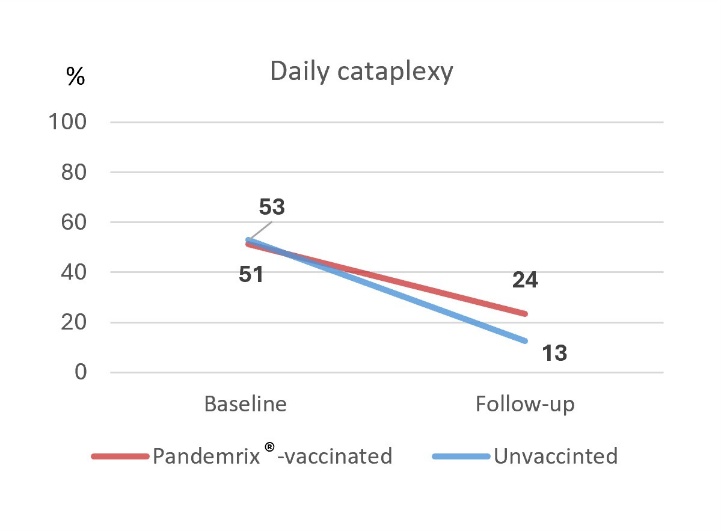

NT1= narcolepsy type 1. The frequency of cataplexy is dichotomized: daily vs. less than daily and never.

Frequency of cataplexy is missing for one individual at baseline, one individual at follow-up and one individual at both baseline and follow-up; all Pandemrix®-vaccinated.
