## Supplemental Figure 1 for "Course and severity of post-H1N1 narcolepsy type 1: a long-term prospective cohort study"

**Figure S1a-c: Frequency of core narcolepsy symptoms in Pandemrix®-vaccinated and unvaccinated NT1 individuals at baseline and follow-up**

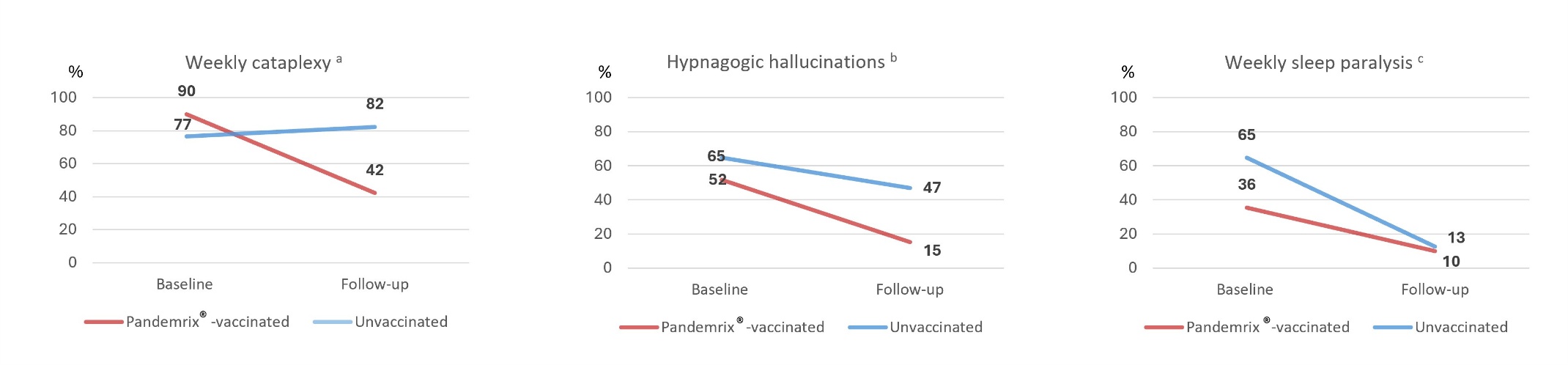

NT1= narcolepsy type 1. The frequency of core narcolepsy symptoms is dichotomized: weekly or more often vs. less than weekly and never.

^a^ Frequency of cataplexy is missing for one individual at baseline, one individual at follow-up and one individual at both baseline and follow-up; all Pandemrix®-vaccinated. ^b^ Frequency of hypnagogic hallucinations is missing for one individual at baseline and two individuals at follow-up; all Pandemrix®-vaccinated. ^c^ Frequency of sleep paralysis is missing for three Pandemrix®-vaccinated individuals at baseline and four Pandemrix®-vaccinated individuals and one unvaccinated individual at follow-up
