## Supplemental Table 4 for "Course and severity of post-H1N1 narcolepsy type 1: a long-term prospective cohort study"

**Table S4: Demographic, laboratory, and medication characteristics of Pandemrix®-vaccinated and unvaccinated individuals**

|  | Pandemrix^®^- vaccinated n=113 | Unvaccinated n=17 | p |
| --- | --- | --- | --- |
| Sex (female) | 71 (62.8 %) | 12 (70.6 %) | 0.535 |
| Age at disease onset (years), median (IQR) | 12.7 (9.9-16.6) | 15.4 (12.5 – 25.6) | 0.140 |
| Diagnostic delay (years), median (IQR) | 2.7 (1.6 – 4.3) | 3.3 (1.2 – 4.8) | 0.833 |
| Age at baseline (years), median (IQR) | 18.3 (15.3 – 24.4) | 20.2 (16.8-30.9) | 0.425 |
| Disease duration at baseline (years), median (IQR) | 6.2 (5.3 -7.6) | 4.8 (4.0 – 7.3) | 0.042* |
| Age at follow-up (years), median (IQR) | 24.0 (21.0 – 30.3) | 26.2 (22.7 – 33.5) | 0.332 |
| Disease duration at follow-up (years), median (IQR) | 12.5 (11.7 – 12.8) | 11.0 (10.2 – 11.6) | 0.003** |
| Follow-up time (years), median (IQR) | 5.7 (4.4-6.5) | 5.5 (4.8 – 6.8) | 0.866 |
| Undetectable hypocretin (hcrt-1 <40 pg/ml) (yes), *n* (%) | 72 (67.9 %) ^a^ | 11 (64.7 %) | 0.792 |
| Medication at follow-up (yes), *n* (%) | 108 (95.6 %) | 14 (82.4 %) | 0.069 |
| Sodium oxybate(yes), *n* (%) | 57 (50.4 %) | 6 (35.3 %) | 0.244 |
| Wake-promoting medication (yes), *n* (%) | 98 (86.7 %) | 13 (76.5 %) | 0.219 |
| Antidepressive medication (yes), *n* (%) | 41 (36.3 %) | 4 (23.5 %) | 0.415 |

IQR= interquartile range.

^*^*p*-values ≤ 0.05. ^**^*p*-values ≤ 0.01. ^***^*p*-values ≤ 0.001.

^a^ CSF-hypocretin-1 level was not measured in four vaccinated individuals and only reported as “low” (i.e. <150 pg/ml) in three vaccinated individuals.
