## Supplemental Table 3 for "Course and severity of post-H1N1 narcolepsy type 1: a long-term prospective cohort study"

**Table S3: Demographic, laboratory, and medication characteristics of NT1 individuals with and without new core NT1 symptoms at follow-up**

|  | Individual with new symptoms ^a^  n=19 | Individuals without new symptoms n=111 | p |
| --- | --- | --- | --- |
| Sex (female), *n* (%) | 13 (68.4 %) | 70 (63.1 %) | 0.202 |
| Age at disease onset (years), median (IQR) | 13.0 (10.8 – 36.5) | 12.9 (9.9 – 18.5) | 0.401 |
| Diagnostic delay (years), median (IQR) | 2.9 (1.2 – 3.8) | 2.8 (1.6 -4.8) | 0.587 |
| Age at baseline (years), median (IQR) | 18.7 (13.9-44.3) | 18.6 (15.4-25.8) | 0.950 |
| Disease duration at baseline (years), median (IQR) | 5.5 (2.5 – 6.9) | 6.3 (5.2-7.7) | 0.041* |
| Age at follow-up (years), median (IQR) | 24.8 (19.7 – 48.7) | 24.1 (21.0 - 30.9) | 0.810 |
| Disease duration at follow-up (years), median (IQR) | 12.1 (8.6 – 12.8) | 12.4 (11.2 – 12.8) | 0.262 |
| Follow-up time (years), median (IQR) | 5.9 (5.2 - 6.9) | 5.6 (4.3 – 6.5) | 0.152 |
| Pandemrix^®^-vaccinated (yes), *n* (%) | 15 (79.0 %) | 98 (88.3 %) | 0.266 |
| Undetectable hypocretin (hcrt-1 <40 pg/ml) (yes), *n* (%) | 13 (72.2 %) ^b^ | 70 (66.6 %) ^c^ | 0.642 |
| Medication at follow-up (yes), *n* (%) | 18 (94.8 %) | 104 (93.7 %) | 0.861 |
| Sodium oxybate (yes), *n* (%) | 5 (26.3 %) | 58 (52.3 %) | 0.037* |
| Wake-promoting medication (yes), *n* (%) | 18 (94.7 %) | 93 (83.8 %) | 0.212 |
| Antidepressive medication (yes), *n* (%) | 6 (31.6 %) | 39 (35.1 %) | 0.091 |

NT1= narcolepsy type 1. IQR= interquartile range.

^*^*p*-values ≤ 0.05. ^**^*p*-values ≤ 0.01. ^***^*p*-values ≤ 0.001.

^a^ Some of these individuals developed more than one core narcolepsy symptom during the follow-up period. ^b^CSF-hypocretin-1 level was not measured in one individual. ^c^ CSF-hypocretin-1 level was missing in 6 individuals: not measured in three individuals; hypocretin-1 level only reported as “low” (i.e. <150 pg/ml) in three individuals.
