## Supplemental Table 2 for "Course and severity of post-H1N1 narcolepsy type 1: a long-term prospective cohort study"

**Table S2: Associations between phenotype and BMI severity at baseline and follow-up**

|  | BMI at baseline  Children ^a^ Adult | | | | BMI at follow-up  Children ^a^ Adult | | | |
| --- | --- | --- | --- | --- | --- | --- | --- | --- |
|  | β (95% CI) | p | β (95% CI) | p | β (95% CI) | p | β (95% CI) | p |
| Sex (female)  (woman) |  |  | −3.41 (−6.36, −0.46) | 0.024* |  |  | −1.72 (−5.03, 1.58) | 0.301 |
| Age at inclusion (years) |  |  | 0.07 (−0.04, 0.18) | 0.183 |  |  | −0.06 (−0.20, 0.08) | 0.411 |
| ESS-score (years) | 0.09 (0.01, 0.17) | 0.033* | 0.50 (0.18, 0.82) | 0.002** | 0.04 (−0.06, 0.14) | 0.399 | 0.09 (−0.25, 0.44) | 0.592 |
| Daily cataplexy ^b^ (yes) | 0.14 (−0.44, 0.72) | 0.632 | −0.97 (−3.62, 1.67) | 0.465 | 0.14 (−0.98, 1.26) | 0.807 | 1.91 (−2.89, 6.72) | 0.429 |
| Mean MSLT SL | −0.01 (−0.10, 0.08) | 0.810 | −0.37 (−0.82, 0.08) | 0.106 | −0.04 (−0.14, 0.06) | 0.391 | −0.33 (−0.62, −0.04) | 0.027* |
| PSG SSSI | 0.02 (−0.07, 0.10) | 0.681 | 0.26 (−0.01, 0.52) | 0.059 | 0.00 (−0.08, 0.08) | 0.996 | 0.15 (−0.18, 0.48) | 0.375 |

BMI = body mass index. CI= confidence interval. ESS= Epworth sleepiness scale. MSLT= multiple sleep latency test. SL= sleep latency. PSG = polysomnography. SSSI= sleep stage shift index. AHI = Apnoea–Hypopnea Index.

^*^*p*-values ≤ 0.05. ^**^*p*-values ≤ 0.01. ^***^*p*-values ≤ 0.001.

At both baseline and follow-up, the children group includes individuals with age < 18 years at baseline, and the adult group includes individuals with age ≥ 18 years at baseline. BMI data were missing for one of 58 children at baseline and one of 58 children at follow-up (different individuals missing data). Among adults, BMI data were missing for one of 72 adults at baseline and for two of 72 adults at follow-up (different individuals missing data). ^a^ For children, BMI is assessed using age- and sex-adjusted BMI z-scores according to the International Obesity Task Force [59]; therefore, these covariates are not included in the analysis. ^b^ The frequency of cataplexy is analysed dichotomized: daily versus less than daily and never.
