## Supplemental Table 1 for "Course and severity of post-H1N1 narcolepsy type 1: a long-term prospective cohort study"

**Table S1: Predictors for daily cataplexy at baseline and follow-up**

|  | Daily cataplexy at baseline ^a^ | | Daily cataplexy at follow-up ^a^ | |
| --- | --- | --- | --- | --- |
|  | OR (95 % CI) | p | OR (95 % CI) | p |
| Sex (female) | 4.24 (1.8, 10.0) | 0.001*** | 1.62 (0.5, 5.4) | 0.439 |
| Age at inclusion (years) | 0.99 (1.0, 1.0) | 0.635 | 1.00 (1.0, 1.0) | 0.878 |
| Disease duration (years) | 0.86 (0.7, 1.0) | 0.132 | 0.83 (0.6, 1.1) | 0.142 |
| Hypocretin <40 pg/ml (yes) | 1.43 (0.6, 3.3) | 0.392 | 2.22 (0.6, 8.5) | 0.247 |
| Pandemrix^®^-vaccinated (yes) | 1.22 (0.4, 3.7) | 0.718 | 0.53 (0.1, 2.0) | 0.348 |

OR= odds ratio. CI= confidence interval.

^*^*p*-values ≤ 0.05. ^**^*p*-values ≤ 0.01. ^***^*p*-values ≤ 0.001.

^a^ Frequency of cataplexy is analysed dichotomized: daily versus less than daily or never.
